# Liquid-Dendrite Spiking Neural Network for Edge Devices: A 130 K-Parameter, 535 KB Model for Time-Domain Epileptic Seizure Detection

**DOI:** 10.1101/2025.02.09.25321975

**Authors:** Luis Fernando Herbozo Contreras, Leping Yu, Zhaojing Huang, Armin Nikpour, Omid Kavehei

## Abstract

Epilepsy is a significant global health issue, requiring dependable diagnostic tools like scalp encephalogram (scalp-EEG), sub-scalp EEG, and intracranial EEG (iEEG) for precise seizure detection and treatment. AI has emerged as a powerful tool in this domain, offering the potential for real-time, responsive monitoring. Traditional methods often rely on feature extraction techniques like Short-Time Fourier Transform (STFT), which can increase power consumption, making them less suitable for deployment on edge devices. While large models can improve accuracy without STFT, their size also limits their practicality for edge applications. This study introduces Liquid-Dendrite, a novel bio-inspired model for seizure detection, leveraging Liquid-Time Constant Spiking Neurons (LTC-SN) and dendrites spiking neurons (dSN) with heterogeneous time-constants. The model comprises two hidden layers with dendritic neurons and one layer of liquid-time constant networks. Our model achieves a memory efficacy of 535 KB with 130 K trainable parameters. The model was tested across the most noteworthy epilepsy datasets for scalp EEG (TUH and CHB-MIT) and iEEG (EPILEPSIAE). Our model demonstrated commendable performance, achieving AUROC scores of 83%, 96%, and 93%, respectively, outperforming some existing models in an energy and memory-efficient way. Moreover, we conducted a robustness test by blacking out EEG channels at the inference stage, where we showed the ability of our network to work with fewer channels. We could deploy our tiny model and perform inference at the edge of the Raspberry Pi 5 without the need for additional quantization. This highlights the potential of Neuro-Inspired AI for efficient, small-scale, and energy-embedded AI systems across different brain modalities.

## 1. Introduction

Epilepsy affects approximately 1% of the world’s population, making it a significant global health issue [1]. This prevalence has driven substantial research aimed at improving epilepsy treatment, mainly by developing technologies such as artificial intelligence (AI) for detecting and predicting seizures. However, due to the unpredictable nature of seizures, long-term brain signal monitoring is required for both diagnosis and research purposes [2]. Traditional scalp electroencephalogram (EEG) presents several limitations, including complex setup procedures, discomfort, and the presence of numerous wires, which make it highly visible and impractical for continuous use [3, 4]. Although ambulatory EEG (aEEG) offers improved portability, the bulkiness and uncomfortable wearability (incl. skin irritations) aspects of surface or scalp EEG headsets often discourage individuals from wearing them consistently [5]. Consequently, there is a need for EEG monitoring solutions that are discreet, user-friendly, and easier to set up. Wearable technologies, such as ear EEG, have been introduced to overcome these challenges as non-invasive techniques that involve placing electrodes within the ear to capture brain activity. While this approach offers significant advantages for long-term monitoring, including enhanced comfort and discreetness stigma [6], the limited number and placement of electrodes inherent to ear EEG systems pose challenges as less information is available than aEEG. Invasive technologies, such as intracranial EEG (iEEG) involves continuously monitoring and has been used in delivering stimulation solely when epileptiform activity is identified as a mean of responsive treatment [7]. Both technologies can benefit from AI at the edge, which can be used on the device for inference/training, reducing power consumption due to data transmission to a cloud server. However, conventional AI systems often demand substantial memory and energy resources, making them unsuitable for applications that involve long-term monitoring [8, 9]. It is essential to develop models to efficiently optimize memory usage and power consumption to address this challenge, enabling deployment on wearable and edge devices.

### 1.1. Background

Inspired by the brain’s structure, Neuromorphic AI contrasts sharply with the traditional von Neumann architecture, where memory and processing are separated, leading to power inefficiencies during data transmission. Neuromorphic chips, however, integrate memory and processing within neurons and synapses, improving speed and energy efficiency and on-device training [10]. While some models utilize Leaky Integrate-and-Fire (LIF) neurons, their efficacy often falls short, partly due to the simplified nature of these models. A promising direction is incorporating biophysical brain models, particularly those that account for dendrites, which are critical for neuronal computation [11]. Researchers have also utilized Adaptive Spiking Recurrent Neural Networks (ASRNNs), which have demonstrated significant potential in this area, offering enhanced computational capabilities with learnable membrane time constants [12, 13].

### 1.2. Prior-work

Researchers are actively advancing AI models for seizure detection. For example, some studies have combined Independent Component Analysis (ICA) and Short Time Fourier Transform (STFT) for preprocessing, followed by ConvLSTM blocks, achieving an impressive AUROC of 0.84 on the largest epilepsy dataset in the United States [14]. Similarly, another approach employed domain adaptation alongside STFT preprocessing, reporting an AUC of 0.75± 0.03 on CHB-MIT database [15]. Despite these advances, such models often require substantial memory and power, limiting their feasibility for edge-device deployment. Epileptic seizure detection has also progressed through integrating Spiking Neural Networks (SNNs) with neuromorphic hardware designed to replicate the brain’s computational strategies for real-time, low-power processing. These systems have demonstrated success with scalp and intracranial EEG data [16]. For instance, a spiking Convolutional Long Short-Term Memory Neural Network (sConvLSTM) achieved high AUROC scores in seizure detection while maintaining nearly 100× –1000 ×power savings compared to traditional methods, with only a 10% reduction in performance [17]. Seizure prediction using neuromorphic computing has also shown promise, with Convolutional SNNs (CSNNs) achieving an AUROC of 0.95 while reducing computational complexity by 98.58% compared to conventional neural networks [18]. Energy-efficient seizure classification has been achieved with CSNNs, consuming as little as 1.28 *µ*J per classification, making them suitable for wearable applications [19]. Previously, we developed a biologically plausible algorithm for seizure detection using Spiking Neural Networks (SNNs), enabling AI-driven electroceuticals at the edge [20]. This work introduced a Liquid-Time Constant (LTC) model to optimize computational efficiency while improving accuracy [20]. Additionally, we proposed a time-domain, energy-efficient seizure detection system based on dendritic Leaky Integrate and Fire neuron (dLIF), avoiding traditional, computationally intensive techniques like Short-Time Fourier Transform (STFT) or Fast Fourier Transform (FFT), demonstrating robustness across various brain modalities, and achieving low-latency inference with minimal computational requirements in real-world applications [21]. However, no previous studies have been able to link different time constants within a spiking neural network.

### 1.3. Novelty and Significance

1. This is the first study incorporating heterogeneous and liquid-time constants within spiking neural networks.
2. The model significantly reduces computational requirements, with a memory footprint of 535 KB and 130 K trainable parameters.
3. The model demonstrates reliable performance across different brain-signal modalities.
4. The model remains robust even with missing data in channels.
5. The approach ensures inference on edge devices with a latency of 0.81 seconds per batch without additional quantization.

## 2. Datasets

In this study, we utilized 3 datasets: the Temple University Hospital (TUH) Corpus scalp-EEG, the Children’s Hospital Boston (CHB-MIT) dataset and the intracranial-EEG EPILEPSIAE dataset, which can be found in Fig. 2.

### 2.1. Scalp-EEG

#### 2.1.1. TUH

The Temple University Hospital (TUH) Corpus is the largest scalp-EEG dataset available globally, comprising recordings from a diverse cohort of 642 patients (315 male and 327 female), ranging in age from 1 to 90 years. This dataset contains 3,044 seizures with a mean seizure duration of 74.4 seconds, recorded across 19 channels using a scalp EEG modality. The dataset is structured into training and validation splits, with 592 sessions allocated for training and 50 for validation, making it an extensive and balanced resource for developing and validating machine learning models.

#### 2.1.2. CHB-MIT

The Children’s Hospital Boston-MIT (CHB-MIT) dataset is a widely used scalp-EEG dataset primarily focused on pediatric patients. We selected 17 patients, which aligns with the baseline results for comparison. It contains recordings of 3 males and 14 females, ranging from 1.5 to 22 years. The dataset includes 160 seizures with an average duration of 92.5 seconds. Recordings were made using a scalp EEG modality across 22 channels, all sampled at 256 Hz.

### 2.2. Intracranial-EEG

#### 2.2.1. EPILEPSIAE

We trained and tested the model using data from 14 patients, as the datasets differ in montages and the number of channels across the EPILEPSIAE dataset, which ranges from 30 to 114 electrodes and has a sampling rate of 256 Hz. The EPILEPSIAE dataset includes high-quality, long-term EEG, intracranial EEG, and simultaneously recorded ECG data. Invasive recordings are obtained from patients using implanted strips, grids, or depth electrodes (stereo-EEG). The dataset contains extensive interictal periods (background data between seizures), making it a more realistic representation of iEEG data and ensuring that our false negatives are highly reliable.

## 3. Methods

### 3.1. Pre-processing

We applied Independent Component Analysis (ICA) to eliminate artifacts from scalp EEG signals. Initially, the EEG data were divided into 12-second segments, and the ICA algorithm was used to decompose them into 19 independent components through Blind Source Separation (BSS). ICA facilitates the decomposition of EEG signals into statistically independent components, as described by Equ. (1):

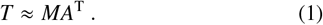

In this equation, *T* represents the EEG data, *M* denotes the time-related information, and *A* corresponds to the weight matrix for the topographic maps. Pearson correlation was utilized to identify independent components highly correlated with eye movement, which was detected using the ‘FP1’ and ‘FP2’ EEG channels. The identified elements associated with eye movement were removed, yielding EEG signals free of such artifacts. Following this, we removed powerline noise from the MNE package from the TUH and CHB-MIT datasets using a notch filter at 60 Hz.

### 3.2. Spiking Neural Network

The proposed method is modeled as an adaptive spiking recurrent neural network incorporating dendrites with heterogeneous time-constants and liquid-time-constant membrane potentials that are inherently learnable systems.

#### 3.2.1. Dendrite Spiking Neuron Structure

The dLIF neuron model improves upon the traditional LIF-based spiking neuron by incorporating multi-timescale memory within the dendrites. This enhancement is described by Equ. (2), Equ. (3), and Equ. (4), where *u* represents the membrane potential of the soma, *β* represents the timing factor, R denotes the membrane resistance, *d* represents the dendritic branch index, and u_th_ represents the firing threshold [22].

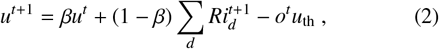

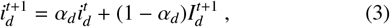

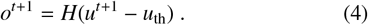

The Heaviside function *H*(·) governs the spiking activity. The synaptic input on the dendritic branch combines feedforward and recurrent inputs [22].

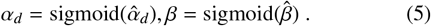

To avoid negative timing factors in Equation (5), *α* and *β* should be restricted within values to 0 and 1, achieved by a sigmoid function.

#### 3.2.2. Liquid Time Constant Spiking Neuron Structure

These time constants are determined differently depending on the type of network being used [13]. Equations (6) and (7) integrate the LTC-SN behavior. The scaling function *σ*() is essential to ensure smooth transitions during learning. Equ. (8) defines *θ*_*t*_ as an adaptive threshold, which helps simulate the realistic adaptive behavior observed in biological spiking neural networks. Equ. (9) and Equ. (10) describe how the membrane potential and spiking rate are regulated by *τ*_*m*_, while Equation (11) defines the resetting of the membrane potential after a spike. Fig. 1 shows the liquid-dendrite neural architecture.

**Figure 1:**
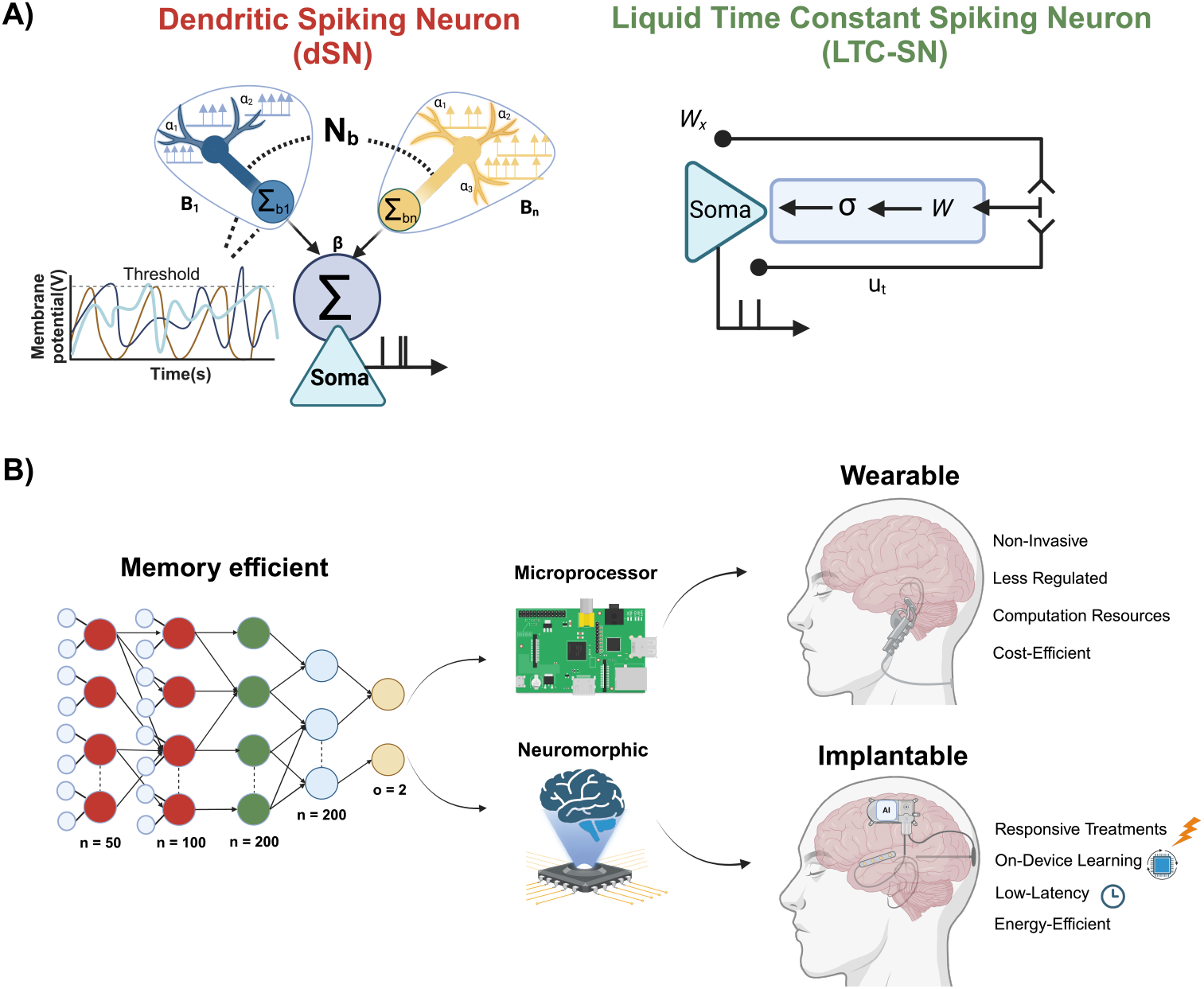
Novel Architecture Framework. Our model features two distinct Spiking Neurons based on dLIF and LTC-SN with learnable parameters. Combining both networks provides a multiple-timescale heterogeneity system for time-series seizure detection (A). Our model consists of two layers of Dendrites with hidden neurons of 50 and 100, respectively, in each layer. The Liquid-Neural Model consists of 200 hidden neurons, connected to 200 dense layers and 2 output classes for both seizure and non-seizure (B), with potential for both wearable and implantable devices.

**Figure 2:**
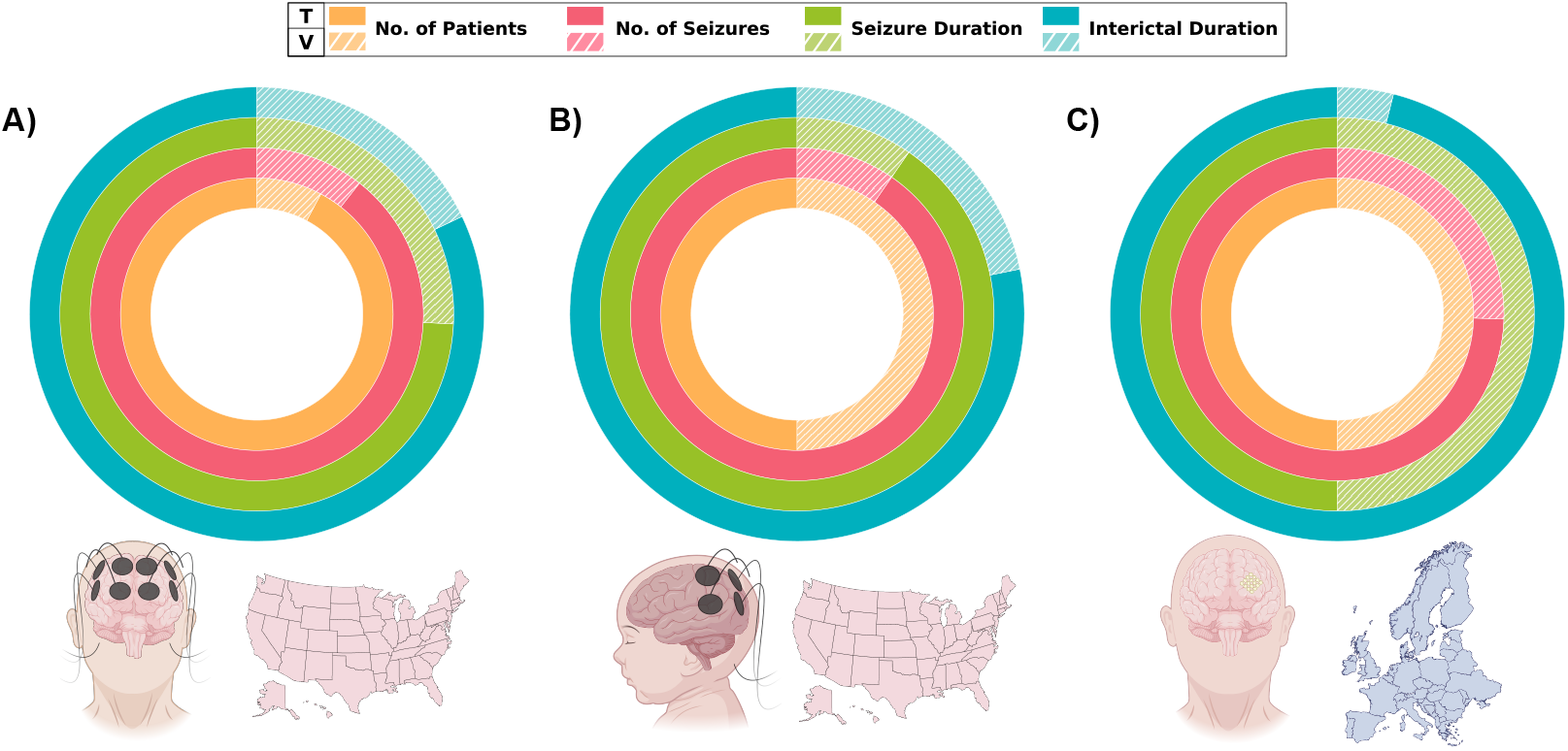
Datasets used in this study. TUH, a large scalp-EEG dataset from adult patients in the USA (A). CHB-MIT, a pediatric scalp-EEG dataset from the USA (B). iEEG-EPILEPSIAE, the largest long-term intracranial-EEG dataset in Europe (C). T: Training, V: Validation.

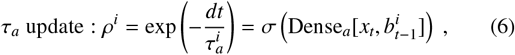

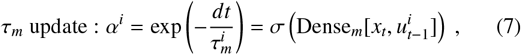

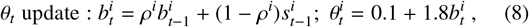

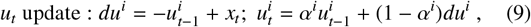

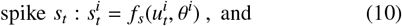

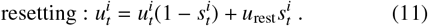

### 3.3. Performance Metrics

The dataset used for model training and evaluation includes labeled data for each 12-second time window. A seizure is identified when the model accurately predicts an ictal event (class 1) based on the ground truth, while non-seizure events are labeled as class 0. Seizure datasets are often imbalanced, with a much higher number of non-seizure data points, as seizures are rare and brief. Over a day, seizure occurrences represent only a small fraction of the data within a 12-second window. Using traditional metrics like accuracy is misleading in this scenario, as it tends to favor the majority class and does not effectively measure the model’s ability to detect seizures, which is the primary goal. For this reason, we rely on the Area Under the Receiver Operating Characteristic Curve (AUC-ROC), which evaluates both sensitivity and specificity in a threshold-independent manner.

### 3.4. Implementation Details

We trained the model on both scalp-EEG and iEEG data for 200 epochs, with a learning rate of 0.001 for the base parameters of the dendrite model. Adam was selected as the optimizer. The time membrane constants for the dendrite layers were initialized uniformly. We employed Xavier uniform distribution for the liquid model to initialize the learnable weights. To distinguish between the seizure (1) and non-seizure (0) classes, we used Mean Squared Error (MSE) as the loss function. Additionally, in our experiments with the scalp-EEG dataset, we reduced the sampling rate to challenge our model with lower-resolution data. This modification significantly reduced memory computation and enabled training to be approximately twice as fast. Training parameters for each dataset can be found in Table 1, and the training algorithm based on Back-Propagation through time is detailed in Algorithm. 1

**Table 1:**
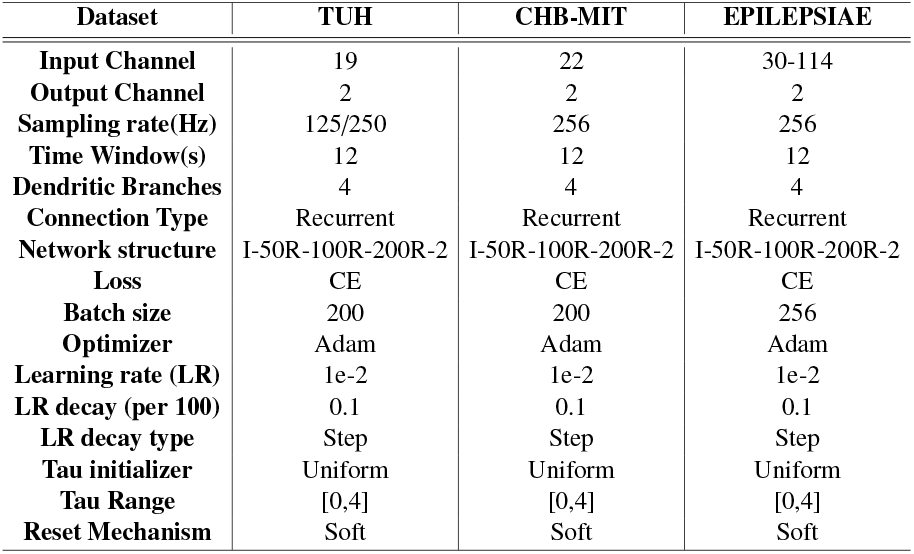
Summary of hyperparameters settings for each dataset.

| Dataset | TUH | CHB-MIT | EPILEPSIAE |
| --- | --- | --- | --- |
| Input Channel | 19 | 22 | 30-114 |
| Output Channel | 2 | 2 | 2 |
| Sampling rate(Hz) | 125/250 | 256 | 256 |
| Time Window(s) | 12 | 12 | 12 |
| Dendritic Branches | 4 | 4 | 4 |
| Connection Type | Recurrent | Recurrent | Recurrent |
| Network structure | I-50R-100R-200R-2 | I-50R-100R-200R-2 | I-50R-100R-200R-2 |
| Loss | CE | CE | CE |
| Batch size | 200 | 200 | 256 |
| Optimizer | Adam | Adam | Adam |
| Learning rate (LR) | 1e-2 | 1e-2 | 1e-2 |
| LR decay (per 100) | 0.1 | 0.1 | 0.1 |
| LR decay type | Step | Step | Step |
| Tau initializer | Uniform | Uniform | Uniform |
| Tau Range | [0,4] | [0,4] | [0,4] |
| Reset Mechanism | Soft | Soft | Soft |

#### Algorithm 1

Training Liquid-Dendrite with BPTT

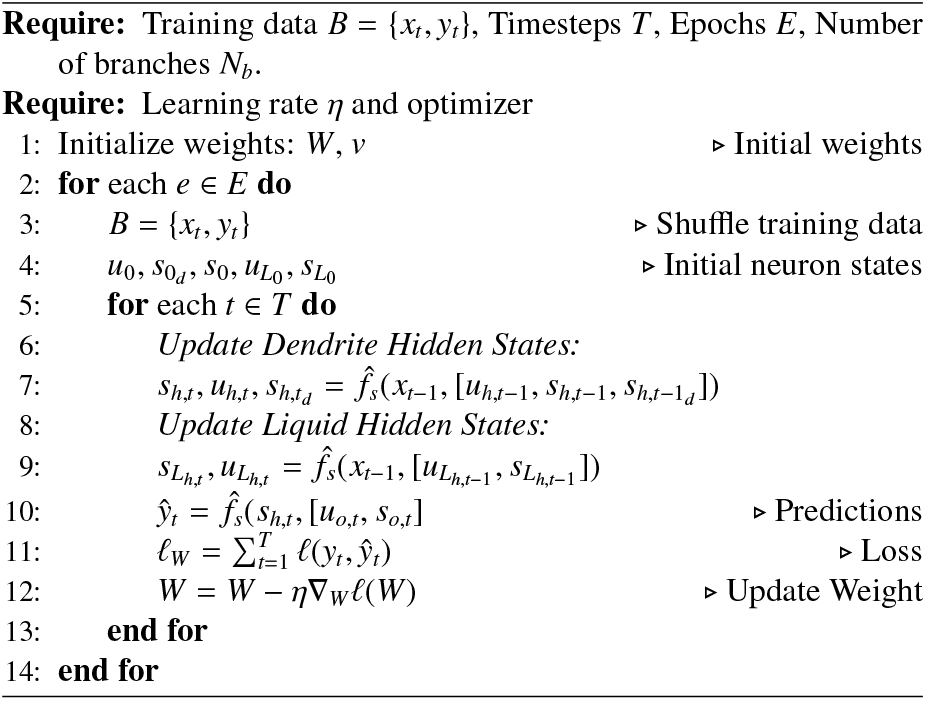

## 4. Results

### 4.1. Neural Network Analysis

#### 4.1.1. Timing Factors

We decided to support our findings by extracting crucial information through the analysis of both the heterogeneous timing factors of the dendrite branches and the liquid timing factors. In Fig. 3, the first 2 layers, which are based on the dendrite spiking neurons, show a higher density at higher timing factors, which are related to heterogeneous system [23, 22]. Interestingly, the liquid time factors show a normal distribution around 0.5. This could be due to the nature of the dense functions governing their value while training.

**Figure 3:**
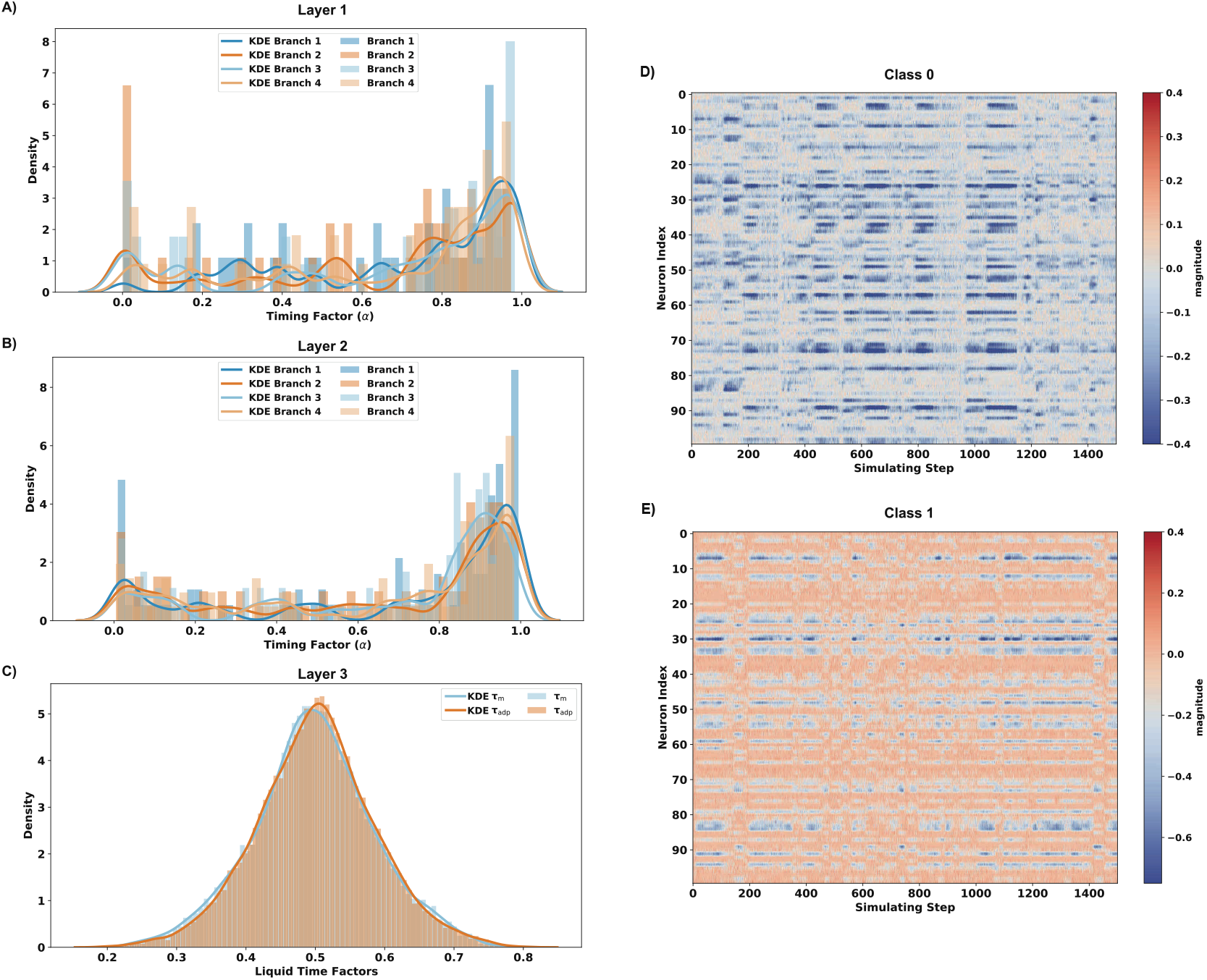
Analysis of learned timing factors and membrane potentials. Our neural network is composed of two layers which are based on dendrites, with the timing factor of each branch showing a higher consolidation of higher timing factors for both layer 1(A) and layer 2(B). Layer 3 has 2 liquid time factors, and overall density follows a normal distribution as both factors are sigmoid (C). Membrane potentials before the last 2 neurons for a random class 0 (D) and random class 1 different (E) show distinct accumulations, which are inherently related to the dynamics of the input data.

#### 4.1.2. Membrane Potential

We extracted the membrane potentials for two random classes. Class 0, which is non-seizure, shows fewer membrane potential voltages, which, in turn, will make the system spike less. While for class 0, which is seizure data, there are more membrane potential activations. One explanation for these findings is that the model is sensitive to identifying seizure data due to higher voltages/amplitude of the input signal. At the same time, the state of class 0 is lower in amplitude than a seizure file.

The simulating steps correspond to 12 seconds of data sampled at 125 Hz.

### 4.2. Scalp-EEG

Our novel architecture achieved an AUROC of **83%** on the TUH scalp-EEG within the time-domain. As depicted in Fig. 4, our model has stable training loss, and metrics for AUPRC and AUROC are also provided. Table 2 shows that our model excels in performance despite using a time domain, spiking network and a very small memory size with fewer parameters and a lower sampling rate. For the CHB-MIT pediatric dataset, we achieved an AUROC of **96%**, which we compared against another state-of-the-art model under the same basis as depicted in Table 3.The number of patients and patient ID reported were selected to provide a direct comparison with previous studies utilizing the same cohort.

**Table 2:**
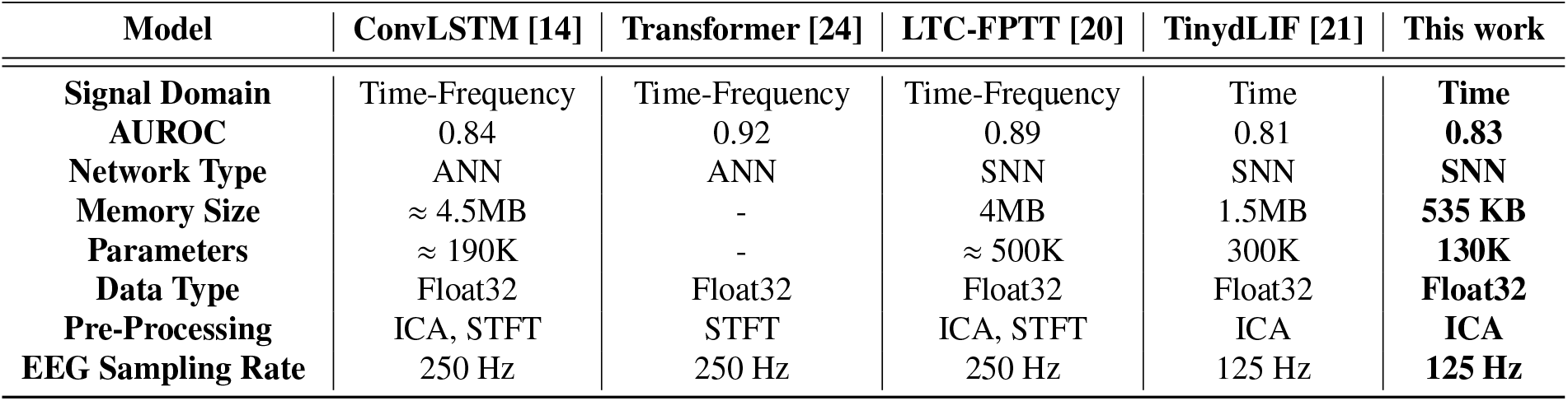
Comparison of performance and metrics of state-of-art models across the TUH dataset.

| Model | ConvLSTM [14] | Transformer [24] | LTC-FPTT [20] | TinydLIF [21] | This work |
| --- | --- | --- | --- | --- | --- |
| **Signal Domain** | Time-Frequency | Time-Frequency | Time-Frequency | Time | **Time** |
| **AUROC** | 0.84 | 0.92 | 0.89 | 0.81 | **0.83** |
| **Network Type** | ANN | ANN | SNN | SNN | **SNN** |
| **Memory Size** | ≈ 4.5MB | - | 4MB | 1.5MB | **535 KB** |
| **Parameters** | ≈ 190K | - | ≈ 500K | 300K | **130K** |
| **Data Type** | Float32 | Float32 | Float32 | Float32 | **Float32** |
| **Pre-Processing** | ICA, STFT | STFT | ICA, STFT | ICA | **ICA** |
| **EEG Sampling Rate** | 250 Hz | 250 Hz | 250 Hz | 125 Hz | **125 Hz** |

**Table 3:** CHB-MIT Results: AUC-ROC comparison of models using time-frequency and time-domain inputs.

| Patient ID | Gender | Time-Frequency Domain |  |  | Time Domain |  |
| --- | --- | --- | --- | --- | --- | --- |
|  |  | CNN [17] | Conv-LSTM [17] | SNN<br>Conv-LSTM [17] | dLIF [21] | dSN-LTC<br>(This work) |
| 1 | F | 100.0 | 100.0 | 96.4 | 98.6 | 100.0 |
| 3 | F | 100.0 | 100.0 | 92.3 | 100.0 | 100.0 |
| 5 | F | 100.0 | 100.0 | 95.3 | 86.0 | 100.0 |
| 6 | F | 94.9 | 93.4 | 88.8 | 93.2 | 88.2 |
| 7 | F | 97.0 | 96.5 | 92.0 | 96.7 | 100.0 |
| 8 | M | 99.4 | 98.2 | 72.5 | 98.3 | 96.2 |
| 9 | F | 99.7 | 100.0 | 96.1 | 98.1 | 91.2 |
| 10 | M | 100.0 | 100.0 | 97.5 | 97.8 | 100.0 |
| 11 | F | 98.8 | 98.9 | 81.2 | 97.7 | 98.3 |
| 15 | M | 99.9 | 99.7 | 73.0 | 95.1 | 81.9 |
| 17 | F | 88.2 | 91.0 | 87.7 | 94.3 | 97.5 |
| 18 | F | 98.5 | 96.9 | 86.0 | 97.3 | 93.7 |
| 19 | F | 95.4 | 99.8 | 95.9 | 92.3 | 99.6 |
| 20 | F | 97.1 | 97.7 | 87.9 | 96.0 | 98.4 |
| 21 | F | 99.3 | 100.0 | 92.5 | 81.4 | 90.1 |
| 22 | F | 100.0 | 100.0 | 96.6 | 90.3 | 97.6 |
| 23 | F | 100.0 | 100.0 | 95.9 | 88.7 | 96.0 |
| **Average** | 3M/14F | 98.13 | 98.36 | 89.9 | 94.2 | 95.8 |

M: Male, F: Female.

**Figure 4:**
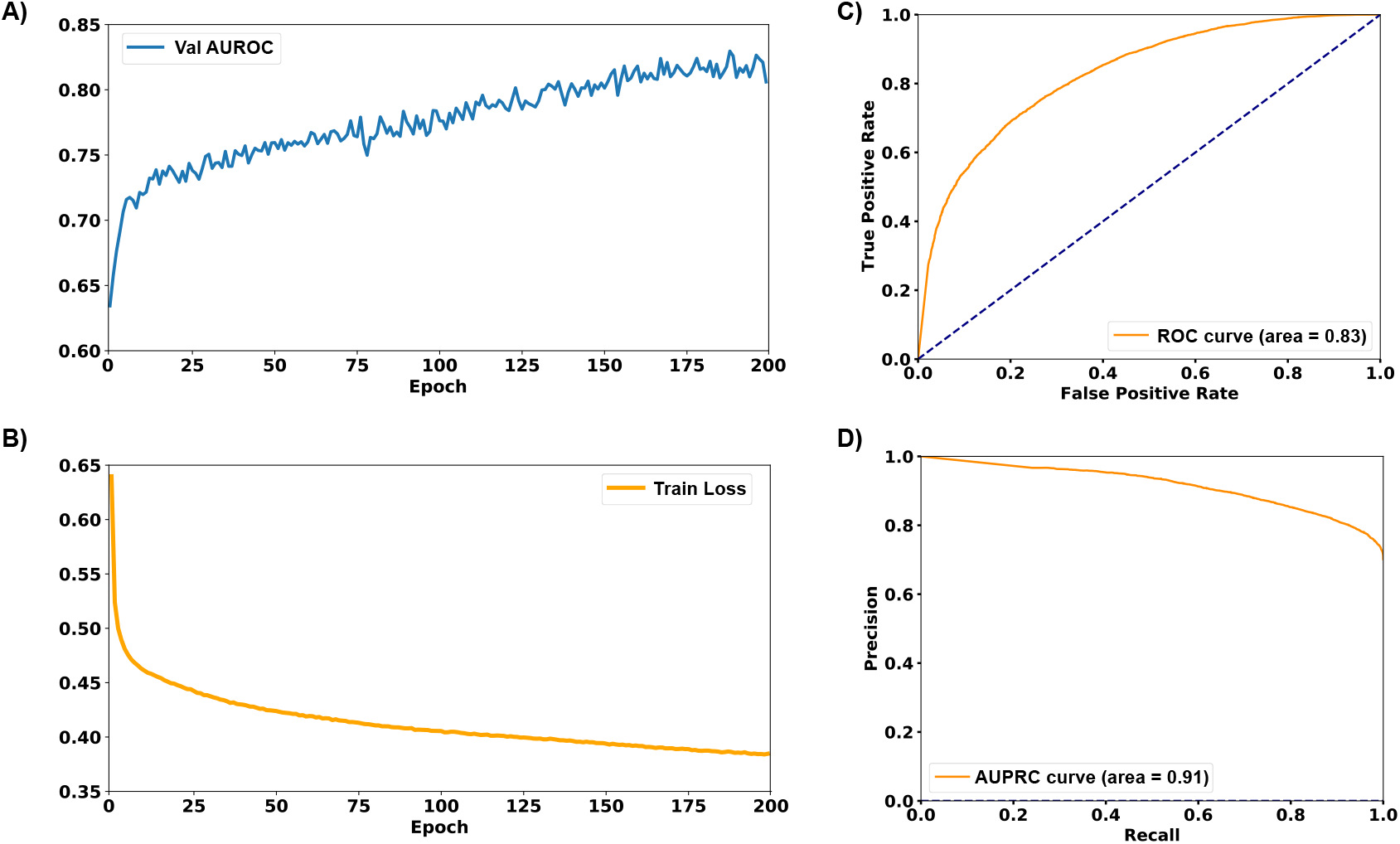
Training and validation results across the TUH epilepsy dataset. Validation AUROC (A) shows an improvement as the training goes on. Train Loss achieves a convergence (B). AUROC (C) and AUPRC (D) curves are presented and show an optimal performance on our network.

### 4.3. Intracranial-EEG Results

When applied to iEEG data, the model achieved an AUROC of **93%**, comparable to larger models that rely on more complex feature extraction techniques and traditional artificial neural networks. Results per patient can be found in Table 4.

**Table 4:**
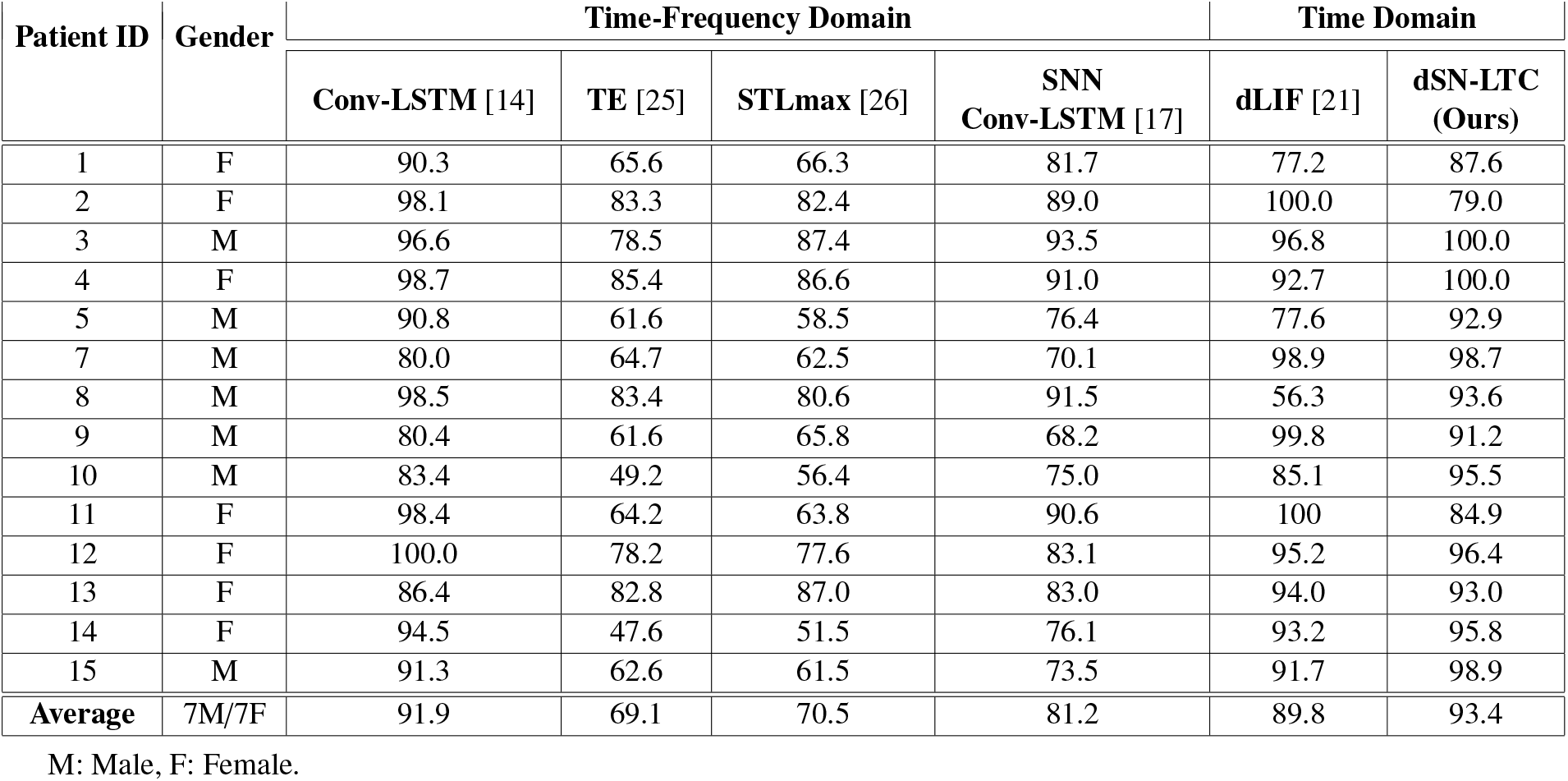
EPILEPSIAE-iEEG Results: AUC-ROC comparison of models using time-frequency and time-domain inputs.

### 4.4. Robustness test

#### 4.4.1. TUH dataset

We experimented to evaluate the robustness of our neural network on the TUH dataset by randomly blacking out between 10% and 90% of the EEG channels during each batch. This approach introduces additional variability and builds confidence in the model’s resilience to channel reduction. The results in Fig. 5 demonstrate that the neural network maintains strong performance even as the number of available channels decreases. Notably, with a 50% reduction in channels, the model achieved an accuracy of approximately 76%. This performance is competitive compared to other neural networks [27, 28, 29], even when utilizing the maximum number of channels.

**Figure 5:**
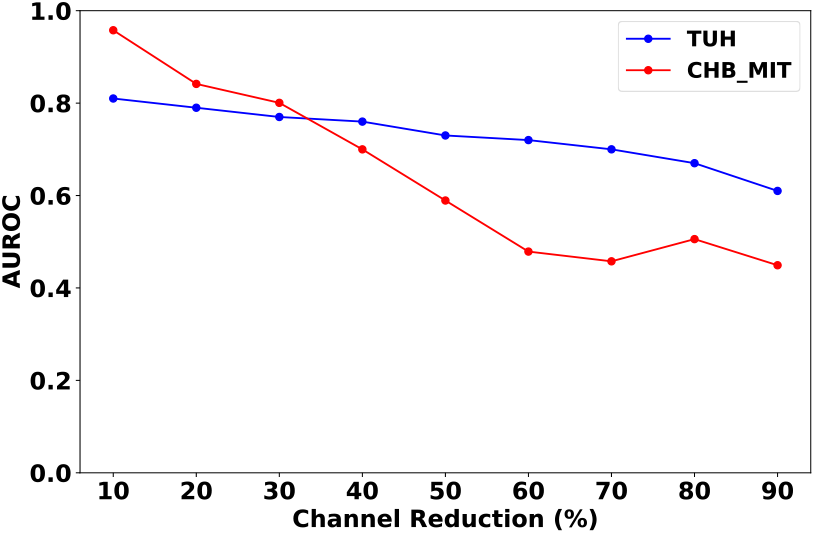
Black-out channels results on the TUH and CHB-MIT dataset.

#### 4.1.2. CHB-MIT dataset

Similarly, we conduced an experiment. Fig. 5 shows the average of all patients in 5 trials. We randomly blackout channels from 10% and 90% to evaluate the performance on the children dataset. Although results are slightly worse than TUH dataset, our model tends to have an acceptable resilience up to a 50% of reduction channels, which highlights the importance of including liquid-time constants and heterogeneous behavior in the neural network.

These findings highlight the advantages of energy-efficient systems where channel dependency is optimized to prioritize only the necessary.

### 4.5. Edge-Inference Raspberry Pi

We performed inference on the edge by utilizing a Raspberry Pi 5. Although this version has a bigger memory, our RAM consumption was limited to 2 GB. We chose Raspberry Pi 5 as it allows the utilization of the latest PyTorch version and Python framework. We measured the inference time for 1 second of EEG data comprising 256 readings sampled at 125 Hz. By averaging the time taken to process all batches of 10,000 EEG signals, we observed an inference time of 0.81 seconds per batch. We recorded an average processing time of 3.1 ms per EEG reading on the fly.

## 5. Limitations

Currently, our method employs Back-Propagation Through Time (BPTT) for training. In future work, we plan to explore Forward Propagation Through Time (FPTT), which will enable the analysis of more time steps with a reduced increase in computational complexity, thereby enhancing efficiency. Our current training approach does not yet extend to addressing out-of-sample generalization tasks. However, the model’s performance on the validation dataset indicates its potential effectiveness in other tasks. Additionally, the training speed of our method prompts us to consider alternative training methodologies that could accelerate the process, which can be enhanced with encoding schemes [30]. Moreover, quantization-aware training can lead to faster inference times [31].

## 6. Conclusion

The proposed method demonstrates efficiency even at a smaller model size, making it well-suited for demanding tasks like seizure detection. We further challenged our model by opting to analyze data purely in the time domain rather than relying on conventional time-frequency domain feature extraction. This approach simplifies the data processing pipeline and offers significant advantages towards an envisioned on-the-edge training. Further, a reduction in the sampling rate didn’t show any decrease in the performance. This methodology can be particularly beneficial for neuromodulation devices, where privacy concerns may arise from transferring sensitive information to cloud servers. By enabling real-time, on-device learning and analysis, our approach can potentially mitigate these concerns while ensuring efficient and effective performance in critical applications.

## Data Availability

The TUH dataset can be freely obtained https://isip.piconepress.com/projects/tuh_eeg/. Access to the EPILEPSIAE dataset requires payment and is accessible https://www.epilepsy-database.eu/. The Children s Hospital Boston dataset is publicly available https://physionet.org/content/chbmit/1.0.0.

https://www.epilepsy-database.eu/

https://physionet.org/content/chbmit/1.0.0

## 7. Code Access

If you have any questions regarding access to the code employed in this research paper, please direct your inquiries to the corresponding author.

## 8. Acknowledgement

Luis Fernando Herbozo Contreras would like to acknowledge the partial support of the Faculty of Engineering Research Scholarship provided by The University of Sydney. Zhaojing Huang would like to acknowledge the support of the Research Training Program (RTP) provided by the Australian Government. The research is supported by the Australian Research Council under Project DP230100019.

## 9. Data Accessibility

The TUH dataset can be freely obtained here. Access to the EPILEPSIAE dataset requires payment and is accessible here. The Children’s Hospital Boston dataset is publicly available here.

